# Mapping the Brain Network of Conduct Disorder: Heterogeneous fMRI findings converge on a Common Brain Circuit

**DOI:** 10.1101/2024.06.02.24308339

**Authors:** Jules R. Dugré, Stéphane Potvin

## Abstract

Conduct disorder (CD) is among the most prevalent and burdensome disorders in early adolescence. Over the past decade, there has been growing interest in identifying reliable and localized neurobiological markers of conduct disorder (CD). However, recent meta-analyses have highlighted the weak reliability of these so-called markers, thereby limiting the ability to draw firm conclusions. Using normative network mapping (598 healthy subjects), we rather sought to investigate whether the heterogeneous findings across studies may map unto a common brain network. A meta-analysis of 38 fMRI studies involving adolescents with a CD (932 cases, 975 controls) was first conducted and showed only a very weak spatial convergence in brain activity alterations in the anterior temporal lobe (5 out of 38 studies). In turn, network mapping revealed that findings across studies show a consistent connectivity pattern across the whole brain, with regional overlap reaching up to 94.7% (36 out of 38 studies). This network was primarily driven by functional connectivity of brainstem nuclei, subcortical structures (i.e., thalamus, ventral striatum), cingulate cortex (i.e., anterior to posterior midcingulate), superior temporal sulcus, and visual cortices. We further describe the neurochemicals and genetic markers of this CD-Network with emphasis on midbrain serotoninergic, dopaminergic and cholinergic projections. Our findings suggest that our understanding of the neurobiological markers of CD could be enhanced by viewing the brain as a complex interconnected system rather than reducing its complexity to a limited number of brain structures. More importantly, this CD-Network may serve as evidence that the various theories of CD can be reconciled rather than seen as conflicting.

## Introduction

Conduct disorder (CD) is generally defined as persistent patterns of severe antisocial behaviors including various aggressive and rule-breaking behaviors ^1^. Its global prevalence has been estimated to be around 5% ^2^ and is considered among the most burdensome psychiatric disorders in early adolescence ^3^. While this disorder continues to be defined exclusively by behavioral criteria, early theories posited that youths with CD may exhibit potential deficits in neuropsychological processes (e.g., verbal and executive functions, ^4^), attentional processes ^5^, reward processing ^6^, conditioning to distressing cues ^7, 8^, and under-arousal, as a marker of fearlessness ^9^ and/or stimulation-seeking ^10, 11^. Indeed, a bulk of psychological research now recognizes that youths with CD exhibit widespread impairments in interpersonal ^12–14^ and cognitive functioning ^15, 16^. In addition, significant progress has been made to identify the potential neurobiological mechanisms underpinning CD. For example, a variety of physiological (i.e., reduced heart rate, low skin conductance), neuroendocrine (i.e., decreased function of the hypothalamic-pituitary-adrenal axis), and neurochemical (i.e., decreased serotoninergic, noradrenergic, and dopaminergic systems) features have been associated to CD ^17–20^. In neuroimaging literature, however, the search for neurobiological markers of CD remains one of the biggest challenges in the field, underscoring both the complexity of brain-behavior relationships and the necessity for further integrative research.

Literature reviews and meta-analyses of adolescents exhibiting severe conduct problems (including those diagnosed with a CD) highlighted aberrant brain activity during a wide range of neurocognitive domains including emotion processing, reinforcement learning, executive functions, and social cognition ^2, 21–24^. Most notable evidence indicate that the amygdala, the orbitofrontal cortex/ventromedial prefrontal cortex (OFC/vmPFC), the anterior insula (aINS) and the anterior cingulate cortex (ACC) may be key regions involved in the pathogenesis of CD ^2, 21^. Indeed, lesion studies in humans also support the crucial role of specific brain regions in the emergence of conduct problems, and potentially CD. For example, Anderson and colleagues ^25^ reported a case in which a patient suffered a trauma to the PFC (frontopolar, vmPFC/mOFC) at an early age and subsequently developing signs of CD including lying, shoplifting, runaway, abusive behaviors and limited prosocial emotions. Others have documented cases involved severe damage to the OFC ^26^ and tumors to the mesial temporal lobe ^27^ which were *causally* linked to emergence of aggressive behaviors during adolescence. In light of these findings, it is unsurprising that these regions have garnered significant attention in neuroimaging research on CD and related disorders ^2, 21, 28–30^. However, findings from fMRI studies reveal a more complex story than what was initially assumed.

Most meta-analyses of fMRI studies in CP/CD have revealed small effect sizes and relatively weak statistical significance across most brain regions ^22–24, 31, 32^. While it is possible that robustness may be altered by the diverse sampling and methodological differences between studies (e.g., clinical heterogeneity, variability in fMRI tasks and statistical analyses, software used, statistical thresholding), it is noteworthy to mention that most regional effects are suspected to be driven primarily by a small subset of studies (≈ 25%) ^33^. As the field of functional neuroimaging struggles with a replicability crisis ^34, 35^, focusing solely on a limited number of brain regions to capture the mechanisms underpinning CD appears untenable. More importantly, this localizationist perspective is susceptible to biological reductionism ^36^ and does not align with current knowledge indicating that across time and conditions, the entire brain operates in *concerto*, through a modular organization of densely interconnected regions ^37, 38^. Indeed, cerebral blood flow and variability of blood oxygen level-dependent (BOLD) signal in local areas are associated with their level of functional connectivity (i.e., node strength/degree centrality) ^39–43^, suggesting that local variability in BOLD signals may reflect information communication ^44^. These findings are coherent with the fact that regions with high degree centrality, especially those involved in cognitively demanding task demands (e.g., reading, memory, inhibition), may demand more energy given their degree of signalling (e.g., supplies in oxygen, glucose) ^45, 46^. Averaging BOLD signal across task conditions and subjects (study-level) would thus reduce the importance of non-hubs regions (those with fewer connections), which are nevertheless essential for our understanding of psychopathologies. Recently, network mapping approaches have been carried out to address limitations generated by localizationist perspectives. For example, Darby and colleagues (2019) showed that heterogeneous locations of brain lesions temporally linked to antisocial behaviors may map onto a common functional connectivity network. These approaches were subsequently adopted in meta-analytic contexts, which were able to reveal, despite the use of heterogeneous peak coordinates, a specific whole-brain network for substance use disorder ^47^, unipolar depression ^48^, and emotion processing ^49^. Adopting a more distributed perspective of brain functioning, as opposed to reducing complex phenotypes to a limited set of brain regions, may be better suited to enhance our understanding of the neurobiological markers for various disorders and psychopathologies ^50^.

In the current study, we aimed to investigate whether using a distributed approach (i.e., network mapping) may outperform common regional approaches in terms of spatial convergence. More precisely, we conducted activation network mapping to examine whether the heterogeneous brain locations found across 38 fMRI studies of adolescents diagnosed with a CD may map onto a common brain network. We subsequently investigated whether the resulting CD-Network spatially corresponded to specific mental functions (e.g., executive functions, social cognition), neurotransmission systems (e.g., serotoninergic, noradrenergic and dopaminergic), and genetic markers (e.g., MAOA, COMT, SLC6A4) that were previously found to be associated with CD and related disorders ^2^.

## Methods

### Meta-analytic Approaches

A subset of studies derived from the most recent meta-analyses of fMRI studies on youths exhibiting antisocial behaviors was included ^23, 24, 51^. These studies were included if they: 1) included a sample with an average of less than 18 years old, 2) included a sample with at least 60% of the participants with a formal diagnosis of CD according to a clinical interview; 3) included a case-control analytic approach; 4) reported the peak coordinates of the significant group-difference across the whole-brain coordinates. To reduce the impact of multiple experiments per study, we concatenated them to form a study-level map ^52^

First, we conducted a coordinate-based meta-analysis on the included fMRI studies across tasks, irrespectively of the directionality of the effect (increased vs. negative), using the activation likelihood estimation algorithm (see Supplementary Method). Spatial convergence was established by using a threshold of p<0.001 at voxel-level and FWE-p<0.05 at a cluster-level with 5000 permutations, as recommended ^53, 54^. This allowed us to establish how many studies contributed to findings at a regional level.

Second, we conducted an activation network mapping of peak coordinates of the included studies. This method involves normative resting-state data to explore to what extent heterogeneous findings may be linked to a common network ^47, 55^. Briefly, a 4-mm sphere was created around each coordinate per each study to create a study-level mask. Then, we computed the normative functional connectivity map of each study-level mask using resting-state data of 598 healthy children and adolescents (mean age=11.87, s.d.= 2.77, 71.6% boys) from the Autism Brain Imaging Data Exchange datasets, ABIDE-I ^56, 57^ and ABIDE-II ^58^ (see supplementary material).

For each study, the group-level connectivity map was computed by averaging the time course of voxels in the study-level mask to the time course of every other voxel in the brain for each of the 598 healthy subjects. Subsequently, group-level connectivity map (across the 598 subjects) was computed using a voxel-wise one-sample *t*-test. To evaluate replicability across studies, we first conducted Spearman correlation between study-level connectivity maps across voxels. We also investigate the level of replicability between studies by thresholding each study-level connectivity map at *t*>5, binarizing it, and combining them. Indeed, others previously found that in some regions, network approaches achieve replicability up to 100% ^47, 55^. Finally, we conducted a voxel-wise one-sample *t*-test using the unthresholded study-level maps to generate a CD-network map that was more consistent than expected by chance through permutation testing ^59^. Main connectivity hubs of this network were identified using cluster-based Threshold-Free Cluster Enhancement (TFCE) and Family-Wise Error corrections (pFWE<0.05). For a more precise description of the top regions characterizing the CD-Network map, several atlases were used including the Yale Brain Atlas ^60^, Automated anatomical labelling atlas (3^rd^ version, ^61^, Harvard Ascending Arousal Network Atlas ^62^ as well as amygdalar (provided by JuBrain Anatomy Toolbox, ^63^), hypothalamic ^64^ and striatal atlases ^65^. Subanalyses were conducted to explore the effect of resting-state fMRI studies, callous-unemotional traits (CU), comorbid attention-deficit/hyperactivity disorder (ADHD) and medication on our findings.

### Functional Decoding: Mental Functions, Neurotransmission and Gene expression

#### Mental functions and Neurotransmission

We compared the spatial associations between the CD-network and 13 meta-analytic maps of mental functions and 20 whole-brain receptor/transporter density maps (see ^66^) using JuSpace (version 1.4) ^67^.

Whole-brain maps of mental functions were used from a data-driven summary of more than 1,347 neuroimaging meta-analyses aiming to derive an ontology of brain functions ^68^. Functions included affective (i.e., motivation, value-based decision-making), cognitive (i.e., language, cognitive control, multiple demand), memory, attention, social cognition (i.e., face perception, social inference, social representation), somatomotor processes (i.e., auditory, interoception, action) ^68^. Voxelwise spearman correlation was conducted between the input (CD-Network) and target images (Mental functions).

PET/SPECT density maps were distributed across 9 neurotransmitter systems including serotonin (i.e., 5-HT_1A_, 5-HT_1B_, 5-HT_2A_, 5-HT_4_, 5-HT_6_, 5-HTT), dopamine (i.e., D_1_, D_2_, DAT), norepinephrine (i.e., NET), Histamine (i.e., H_3_), acetylcholine (i.e., α4β2, M_1_, VAChT), cannabinoid (i.e., CB_1_), opioid (i.e., MOR, KOR), glutamate (i.e., NMDA, mGluR_5_) and GABA (i.e., GABA_A/BZ_). The mean values of 421 brain regions which included 400 parcels of the 7-Network Schaefer atlas ^69^and additional 21 subcortical and cerebellar regions from the ASEG ^70^ and Buckner Atlas ^71^ were extracted for the two images (i.e., CD-Network and target maps [density]). Partial correlation (Spearman’s rank correlation) adjusting for spatial autocorrelation (i.e., local grey matter probabilities) was then performed between the two sets of parcellated maps. Permutation-based p-values (with 5,000 permutations) were computed and corrected using false discovery rate (FDR).

#### 1.1.1. Gene-Category Enrichment Analyses

We investigated whether the CD-Network was significantly associated with gene expression patterns underlying psychiatric disorders (i.e., DisGeNET, ^72^). This was done with the ABAnnotate toolbox (see ^73^) which aims to perform gene-category enrichment analyses using volumetric maps. Brain-wide gene expression patterns were obtained via the Allen Human Brain Atlas ^74, 75^. For these analyses, the CD-network map and the mRNA expression data for 15,633 genes were parcellated using the same parcellations as mentioned above (400 cortical and 21 subcortical and cerebellar regions). Null maps of the CD-Network (5,000) were generated while preserving spatial autocorrelation. Spearman correlations between the CD-network map, the null maps and all the mRNA expression maps were calculated. Positive-sided p-values were then calculated from the comparisons between the “true” category scores with null distribution and were FDR-corrected. To explore the potential role of specific genes, gene-wise spatial associations were then conducted using top genes that characterized disorders associated with the CD-Network.

## Results

### Activation Likelihood Estimation Meta-analysis

A total of 38 fMRI studies was included in the current meta-analysis which comprised 932 cases versus 975 controls (see Table 1). Mean age was 15.5 years old (SD=1.37) and mainly included males (average of 82.2% per sample). The average rates of CD per sample was 91.8% (SD=12.9). Of the 25 studies that reported prevalence of participants receiving medications (average 18.41% of samples), 11 samples comprised unmedicated participants. the Spatial convergence across these studies revealed a significant peak in the anterior inferior temporal gyrus (x=56, y=-10, z=-22, 728 mm^3^, see Figure 1) which was driven by only 5 studies out of 38 (13.15%).

**Figure 1.**
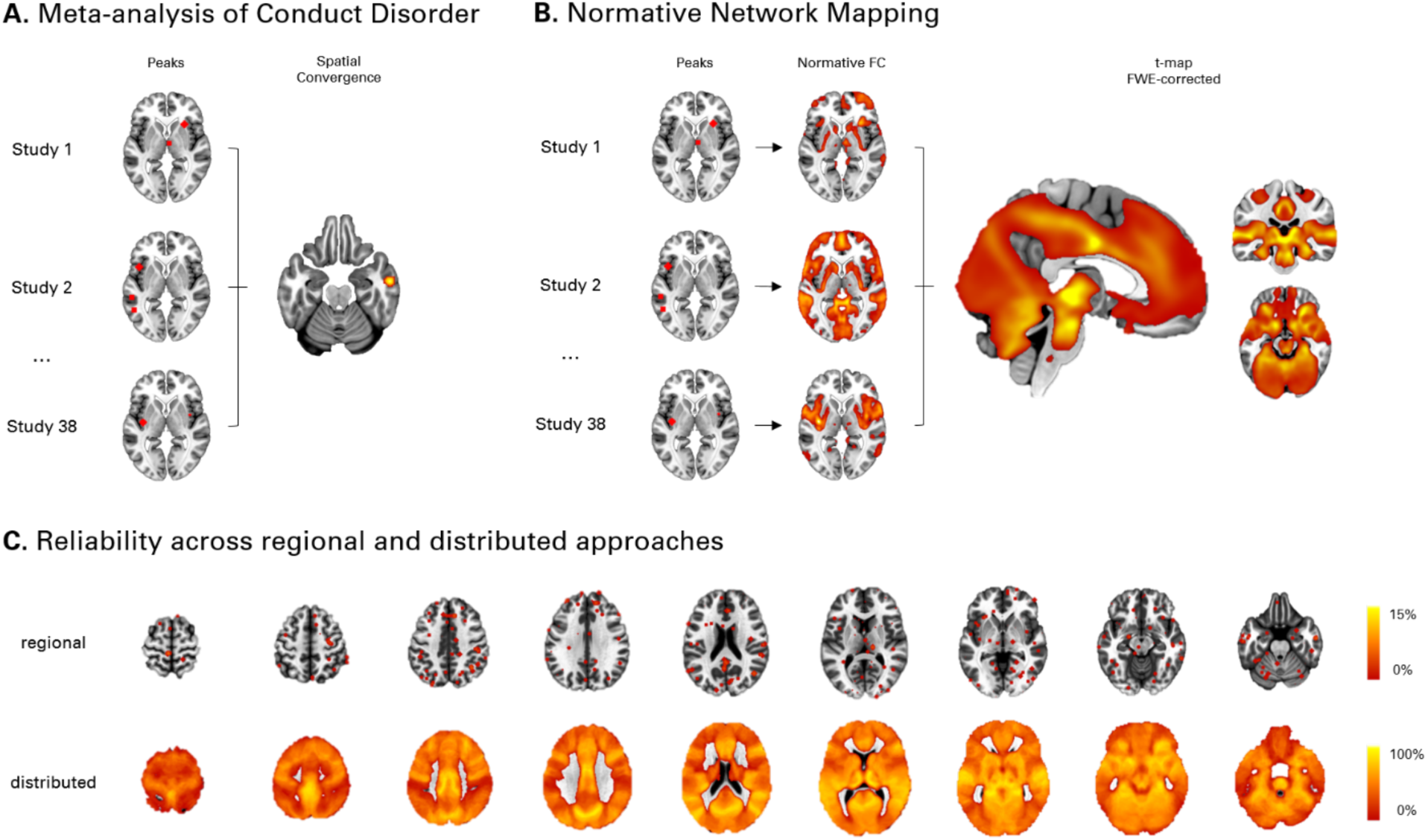
Spatial Convergence across neuroimaging studies on Conduct Disorder. **A.** Activation Likelihood Estimation meta-analysis was conducted on peak coordinates of 38 fMRI studies on Conduct Disorder which revealed a significant convergence anterior inferior temporal gyrus (5 studies out of 38). **B.** Normative Network mapping approach was conducted to identify whether heterogeneous brain locations (peak coordinates) between studies may map onto a common functional connectivity map. Network mapping revealed that study-level maps strongly correlated with each other (average r=.71). **C.** Upper row displays of the overlap between peak coordinates across studies (4 mm sphere) showing low reliability (up to 13.15%). Lower row represents overlap between study-level maps that were binarized (T>|5|) which showed high reliability (up to 97.4% of studies).

**Table 1.** Summary of the Included fMRI Studies on Conduct Disorder (k=38)

| Included Studies<br>(First Author, Year) | Sample characteristics |  |  |  |  |  |  | Task |
| --- | --- | --- | --- | --- | --- | --- | --- | --- |
|  | Controls<br>(N=) | % of<br>Conduct<br>Disorder | Diagnostic<br>Assessment | Cases<br>(N=) | Mean<br>Age | Males<br>(%) | MED<br>(%) |  |
| Aghajani, 2021 <sup>76</sup> | 31 | 100.0% | K-SADS | 19 | 16.4 | 100.0% | 0.0% | Emotional Recognition & Resonance |
| Banich, 2007 <sup>77</sup> | 12 | 75.0% | DISC-IV | 12 | 16.7 | 100.0% | - | Stroop Task |
| Cao 2019 (A) <sup>78</sup> | 30 | 100.0% | SCID-I/P | 36 | 14.3 | 100.0% | - | Rest (ReHo) |
| Cao 2019 (B) <sup>78</sup> | 30 | 100.0% | SCID-I/P | 32 | 14.8 | 100.0% | - | Rest (ReHo) |
| Crowley, 2010 <sup>79</sup> | 20 | 95.0% | DSM-IV CD | 20 | 16.5 | 100.0% | 30.0% | Colorado Balloon Game |
| Crowley et al., 2015 <sup>80</sup> | 20 | 85.4% | DSM-IV | 41 | 16.3 | 49.0% | 34.1% | Decision-Making Behavioral Task |
| Decety 2009 <sup>81</sup> | 8 | 100.0% | DSM-IV | 8 | 16-18 | NA | - | Empathy for Pain |
| Dong, 2016 <sup>82</sup> | 36 | 100.0% | SCID-II | 30 | 15.1 | 100.0% | - | Facial Expression (Pain) |
| Ewbank, 2018 <sup>83</sup> | 25 | 100.0% | K-SADS-PL | 24 | 18.1 | 100.0% | - | Facial Emotional Processing |
| Fairchild, 2014 <sup>84</sup> | 20 | 100.0% | K-SADS | 20 | 17.0 | 0.0% | - | Emotional Face Task |
| Fehlbaum, 2018 <sup>85</sup> | 39 | 100.0% | K-SADS-PL | 39 | 15.9 | 74.0% | 89.7% | Affective Stroop Task |
| Finger, 2011 <sup>86</sup> | 15 | 60.0% | K-SADS-PL | 15 | 14.1 | 60.0% | 46.7% | Probabilistic Reversal Task |
| Gatzke-Kopp, 2009 <sup>87</sup> | 11 | 63.0% | DISC | 19 | 13.6 | 100.0% | 52.6% | Reward Processing |
| Herpertz, 2008 <sup>88</sup> | 22 | 100.0% | K-SADS | 22 | 14.7 | 100.0% | - | Passive-Viewing (Emotional) |
| Hwang, 2016 <sup>89</sup> | 28 | 88.9% | K-SADS | 18 | 14.6 | 56.0% | 27.8% | Affective Stroop Task |
| Hwang 2018 <sup>90</sup> | 29 | 73.7% | K-SADS | 19 | 14.8 | 68.4% | 15.8% | Social & Non-Social Rewards |
| Klapwijk, 2016a <sup>91</sup> | 33 | 100.0% | K-SADS-PL | 23 | 16.6 | 100.0% | 0.0% | Empathic Emotional Face Task |
| Klapwijk, 2016b <sup>92</sup> | 33 | 100.0% | K-SADS-PL | 32 | 16.8 | 100.0% | 0.0% | Dictator Game |
| Lu, 2017 <sup>93</sup> | 18 | 100.0% | K-SADS-PL | 18 | 16.1 | NA | 0.0% | Rest (Short/Long Range Density) |
| Lu, 2020 <sup>94</sup> | 18 | 100.0% | K-SADS-PL | 18 | 17.1 | 100% | 0.0% | Rest (Static/Dynamic ALFF) |
| Lu, 2021 <sup>95</sup> | 18 | 100.0% | K-SADS-PL | 18 | 17.1 | 100.0% | - | Rest (dynamic ReHo) |
| Mathur 2023 <sup>96</sup> | 41 | 63.4% | DSM | 42 | 16.2 | 61.0% | 26.2% | Retaliation Task |
| Menks et al., 2021 <sup>97</sup> | 35 | 100.0% | K-SADS-PL | 23 | 16.7 | 58.3% | - | Emotional Face Processing |
| Passamonti, 2010 <sup>98</sup> | 40 | 100.0% | K-SADS | 27 | 17.1 | 100.0% | - | Emotional Face Task |
| Rachle 2019 <sup>99</sup> | 29 | 86.7% | K-SADS | 30 | 16.3 | 0.0% | - | Effortful Emotion Regulation Task |
| Rubia, 2008 <sup>100</sup> | 20 | 100.0% | Maudsley DI | 13 | 13.0 | 100.0% | 0.0% | Stop Task |
| Rubia, 2009a <sup>101</sup> | 16 | 100.0% | Maudsley DI | 14 | 12.9 | 100.0% | 0.0% | Reward Continuous Performance |
| Rubia, 2009b <sup>102</sup> | 20 | 100.0% | Maudsley DI | 13 | 12.8 | 100.0% | 0.0% | Simon Task |
| Rubia, 2010 <sup>103</sup> | 20 | 100.0% | DSM-IV | 14 | 12.6 | 100.0% | 0.0% | Visual-Spatial Switch Task |
| White, 2012a <sup>104</sup> | 19 | 94.1% | K-SADS | 17 | 15.5 | 76.0% | 11.8% | Eye Gaze Task |
| White, 2012b <sup>105</sup> | 17 | 73.3% | K-SADS | 15 | 15.7 | 80.0% | 26.7% | Emotion-Attention Bars Task |
| White, 2013 <sup>106</sup> | 18 | 85.0% | K-SADS | 20 | 15.2 | 82.0% | 10.0% | Passive Avoidance Task |
| White, 2014 <sup>107</sup> | 15 | 73.3% | K-SADS | 15 | 14.4 | 73.0% | 20.0% | Doors Task |
| White 2016 <sup>108</sup> | 26 | 73.3% | KSADS | 30 | 15.0 | 63.3% | 23.3% | Social Fairness Game |
| Wu 2017 <sup>109</sup> | 28 | 100.0% | SCID-I/P | 28 | 14.8 | 100.0% | 0.0% | Rest (ReHo) |
| Zhou 2015 <sup>110</sup> | 18 | 100.0% | K-SADS-PL | 18 | 16.1 | 100.0% | 0.0% | Rest (ALFF) |
| Zhang, 2015 <sup>111</sup> | 40 | 100.0% | DSM-IV-TR | 29 | 15.1 | 100.0% | - | Go/Stop Task |
| Zhang, 2023 <sup>112</sup> | 77 | 100.0% | DSM | 101 | 15.9 | 56.70% | 45.5% | Passive Avoidance Task |

Note. KSADS = Kiddie Schedule for Affective Disorders and Schizophrenia; DISC = Diagnostic Interview Schedule for Children; ReHo = Regional Homogeneity; ALFF = Amplitude of Low Frequency Fluctuations

### Normative Network Mapping

Next, for each study, we computed the resting-state functional connectivity map using their reported coordinates peaks as seeds (Figure 1B). Using unthresholded study-level maps, a voxelwise one-sample t-test was then conducted to generate an unbiased t-map that is greater than expected by chance. Analyses revealed significant effects in many regions, especially the pons, midbrain, thalamus, ventral striatum, posterior hippocampus, orbitofrontal cortex, cingulate cortices (i.e., posterior midcingulate to perigenual anterior), visual (i.e., V3) and parietal cortices (i.e., anterior superior temporal gyrus to angular gyrus) (Figure 1B, Table 2). Thresholding each study-level t-map at T>|5|, binarizing and summing them revealed that study converge onto a common network with highest voxel replicability reaching up to 94.7% (36 out of 38 studies) in the posterior midcingulate gyrus, posterior hippocampus and thalamus (see Figure 1C and 3A, Table 2). More importantly, using more stringent thresholds than T>|5| (Stubbs et al., 2023) did not impact the findings: T>|6| (overlap<92.1%, r=.98 with T>|5|), T>|7| (overlap<89.5%, r=.96), T>|8| (overlap<89.5%, r=.94), T>|9| (overlap<86.8%, *r*=.91), and T>|10| (overlap<81.5%, *r*=.88). Next, we examined whether our findings might be driven by a particular functional network (Figure 2B). Compared to the rest of the brain, CD-Network was primarily characterized by functional connectivity of the subcortex (Cohen’s *d*=.84), visual (Cohen’s *d*=.80), followed by the default mode network (DMN) (Cohen’s *d*=.67).

**Figure 2.**
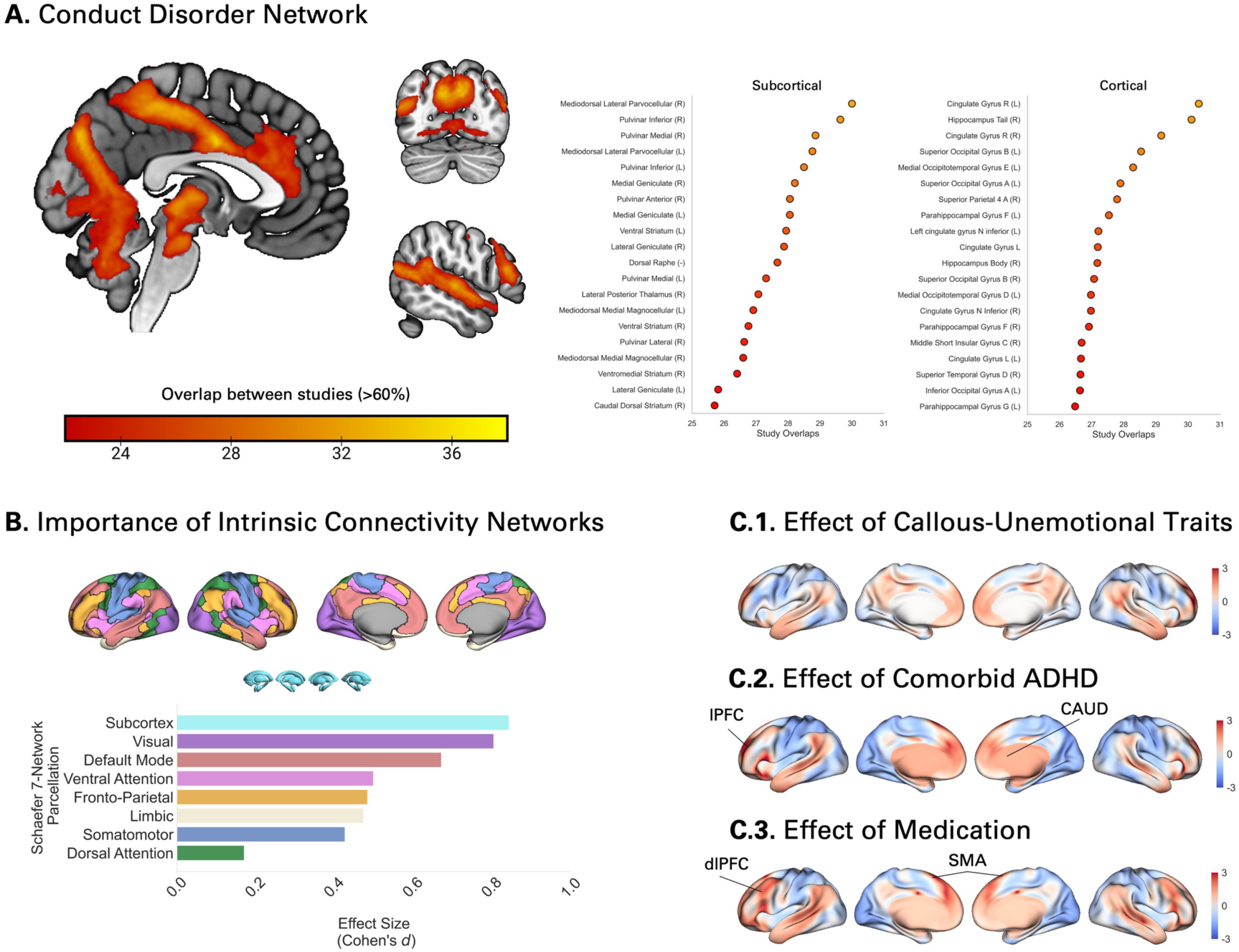
Description of the Conduct Disorder Network (CD-Network). **A.** Figures show top brainstem, subcortical and cortical regions involved in the CD-Network. **B.** Barplot demonstrates the importance of subcortex and somatomotor networks (i.e., Schaefer-7 Networks) in characterizing the CD-Network, compared to the rest of the brain (Cohen’s d). **C.** Subanalyses were conducted to investigate the effect of CU traits, comorbid ADHD and medication. Comorbid ADHD revealed small significant effect in the lateral prefrontal cortex (lPFC) and caudate nucleus (CAUD). Medication level was associated with functional connectivity in the dorsolateral prefrontal cortex (dlPFC), and supplementary motor area (SMA). None of these findings did not survive family-wise error correction (cFWE<0.05)

**Figure 3.**
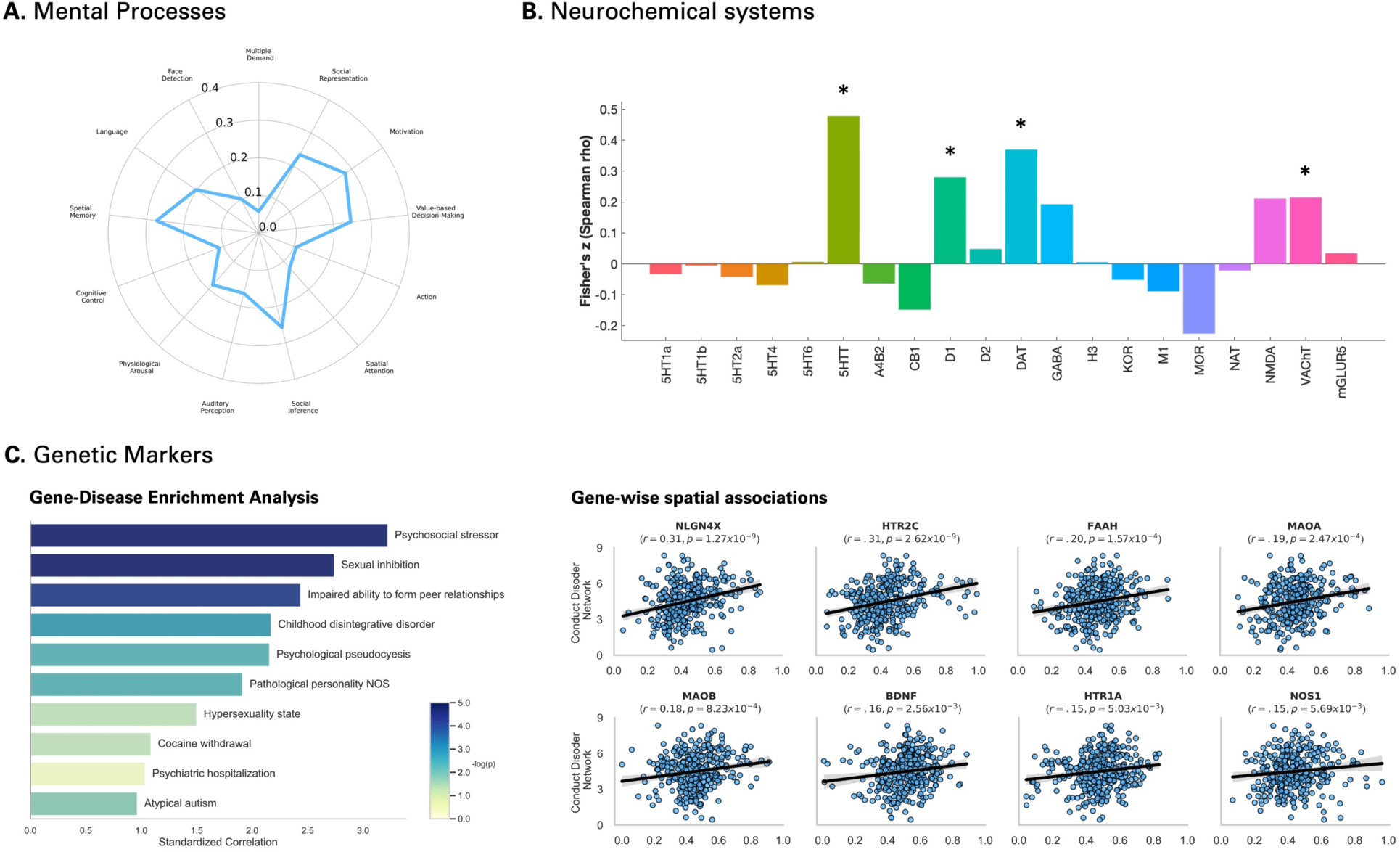
Functional Decoding of the Conduct Disorder Network. **A.** Voxelwise distribution of the CD-Network mainly correlated with motivation, spatial memory, social inference, and value-based decision-making, derived from the brain-behavior ontology of task-based fMRI studies (Dugré et Potvin, 2023). **B.** Top neurotransmission systems that spatially correlated with the CD-Network involved serotoninergic, dopaminergic and cholinergic transporter, and dopamine receptor 1 (Dukart et al., 2020). Asterisks represent statistical significance after correcting for false discovery rate. **C.** Gene-Category Enrichment Analyses were conducted and identified 71 disorders associated with CD-Network. Left panel show the top 10 disorders the correlated the most with CD-Network. Correlations were weighted for the number of gene in categories (i.e., log(number of genes+1), and standardized. Right panel represents gene-wise spatial correlation patterns of the most important genes across the identified disorders.

**Table 2.** Reliability of regional overlap between studies using Normative Network Approach.

| Regions | MNI Coordinates |  |  | Statistics |  |
| --- | --- | --- | --- | --- | --- |
|  | x | y | z | Peak Overlap<br>(out of 38) | t-values |
| pHippocampus | 30 | -32 | -10 | 36 | 10.72 |
| pMCC | -4 | -4 | 34 | 36 | 11.79 |
| Thalamus | 10 | -22 | 12 | 36 | 10.43 |
| Precuneus | -12 | -42 | 44 | 35 | 9.53 |
| Cuneus (V3) | 8 | -74 | 28 | 35 | 6.72 |
| pHippocampus | -22 | -36 | -8 | 35 | 9.11 |
| vPutamen | -16 | 8 | -16 | 34 | 9.88 |
| OFC (Fo3) | 26 | 30 | -14 | 34 | 10.32 |
| ventral Putamen | 28 | 8 | -12 | 33 | 8.78 |
| Superior Temporal Gyrus | 60 | -12 | -2 | 33 | 9.74 |
| Angular Gyrus | 42 | -60 | 18 | 33 | 8.62 |
| IFG (BA 45) | 56 | 32 | 14 | 33 | 9.63 |
| Thalamus | 14 | -28 | -2 | 33 | 9.86 |
| Cerebellum (Dentate Nucleus) | 16 | -60 | -36 | 32 | 10.49 |
| Cerebellum (Dentate Nucleus) | -14 | -60 | -34 | 32 | 9.73 |
| Pons | 4 | -22 | -24 | 32 | 10.29 |
| Amygdala | -30 | 4 | -18 | 32 | 7.89 |
| Superior Temporal Gyrus | -52 | -10 | -4 | 32 | 8.69 |
| Angular Gyrus | -50 | -72 | 16 | 32 | 9.19 |
| posterior Insula | 42 | -24 | 10 | 31 | 7.30 |
| Supramarginal Gyrus | 56 | -42 | 18 | 31 | 6.84 |
| Middle Frontal Gyrus | 26 | 34 | 38 | 30 | 7.06 |

Note. pMCC = posterior Midcingulate Cortex; IFG = Inferior Frontal Gyrus;

Subanalyses were then conducted to investigate the effects of CU traits, comorbid ADHD, and medication on CD-Network (p<0.001, 10 voxels). First, no significant effect was found for the severity of CU traits (see Supplementary Results for median split). Comorbid ADHD revealed a significant effect in the lateral prefrontal cortex (x=-22, y=40, z=14, t=3.8, 125 voxels) and caudate nucleus (x=-16, y=12, z=14, t=3.54, 14 voxels). Medication level was associated with functional connectivity in the dorsolateral prefrontal cortex (x=-30, y=16, z=22, t=5.59, 64 voxels), and supplementary motor area (x=-15, y=-16, z=62, t=3.98, 24 voxels). However, none of these findings did not survive family-wise error correction (cFWE<0.05). Finally, removing resting-state studies (*k*=7) did not impact our findings (voxelwise correlation r=.97).

### Functional Decoding: Mental Functions, Neurotransmission and Gene expression

Given the distributed nature of the CD-Network, we sought to examine whether it spatially corresponded to specific mental functions, receptor density, and genetic markers.

Voxelwise spatial correlations with mental functions maps showed that the CD-Network showed was primarily associated with motivation (r=.28), spatial memory (r=.27), and social inference (r=.26), value-based decision-making (r=.25) and social representation (r=.24). Using PET/SPECT density maps, CD-Network mainly correlated with Serotonin Transporter (5-HTT/SERT)(z’=.48, pFDR<0.05), dopaminergic transporter (DAT)(z’=.37, pFDR<0.05), dopamine receptor D_1_ (z’=.28, pFDR<0.05), and Vesicular Acetylcholine Transporter (VAChT) (z’=.22, pFDR<0.05).

Gene-Category Enrichment Analyses were conducted using postmortem gene expression data to identify psychiatric disorders enriched with genes associated with the CD-Network (DisGenNet). We found that the CD-Network map significantly correlated with gene expression patterns of 71 disorders after correction with 5,000 permutations. Top disorders included Psychosocial stressor (Z=3.22, p=1.3×10^−7^), sexual inhibition (Z=2.74, p=1.1×10^−7^), Impaired ability to form peer relationships (Z=2.44, p=3.2×10^−7^), Childhood disintegrative disorder (Z=2.17, p=2.3×10^−6^), and Psychological pseudocyesis (Z=2.16, p=3.07×10^−6^) (see Supplementary Table 1). More importantly, among these disorders, BDNF (39.4%), MAOA (28.2%), DRD2 (25.4%), COMT (19.7%), HTR1A (19.7%), HTR2A (18.3%), DRD4 (15.5%), and OXTR (15.5%) we the most frequently reported. From these, expression of NLGN4X (rho=.31, p=1.27×10^−9^), HTR2C (rho=.31, p=2.62×10^−9^), FAAH (rho=.20, p=1.57×10^−4^), and MAOA (rho=.28, p=2.47×10^−4^) showed strongest gene-wise spatial correlation with the CD-Network (Figure 3B, Supplementary Table 2).

## Discussion

In the current meta-analysis, we sought to investigate the reliability of neuroimaging findings on CD using both local and diffused approaches. Of the 38 identified studies, spatial convergence in regional brain activity was observed in the inferior temporal gyrus. However, this finding was driven by only 5 studies (13%). In turn, network mapping yielded robust overlap between studies reaching up to 94.7% reliability, particularly in the posterior midcingulate cortex, posterior hippocampus and thalamus. We additionally observed prominent role of functional connectivity in the ascending arousal network (e.g., pontine reticular formation, dorsal raphe), midbrain, ventral striatum, parietal and occipital regions. Furthermore, the CD-Network showed main spatial correlations with socio-affective and reward-related processes, in addition to stronger concordance with density of serotoninergic, dopaminergic, and cholinergic transporters. Finally, gene-category enrichment analyses revealed that the CD-Network was genetically close to psychosocial stress, sexual inhibition and impaired ability to form peer relationships, possibly due to alterations in NLGN4X, HTR2C, FAAH, and MAOA genes expression. Taken together, the CD-Network approach may facilitate the integration of existing theories by providing a robust framework of neural functioning in CD, where distinct mechanisms are functionally intertwined.

Over the past decades, several theorists attempted to describe pathogenesis of CD by proposing seemingly distinct mechanisms that could explain the emergence of antisocial behaviors. Among the most prominent theories, Integrated Emotion systems (IES) posits that the Violence Inhibition Mechanism (VIM) is usually activated by distressing cues, which results in autonomic activity and activation of the threat response system ^7^. Reduced VIM would thus impair stimulus-reinforcement associations between actions that harm others and consequences of such actions (e.g., amygdala, orbito/ventrolateral cortices), and interfere with socialization ^113^. Others rather suggested that CD symptoms may be better explained by alterations in executive functions^4^ and attentional processes ^5^. For example, reduced affective response to threatening stimuli may be due to deficits in shifting the attention of CD individuals to cues that are secondary to their primary focus of attention (Newman, 1987) and/or to an exagerated filtering out cues that are in periphery ^114^. While both emotion-based and cognitive-based accounts mainly rely on distinct neural correlates, they are likely intertwined. For instance, we found a prominent role of brain regions underpinning both reinforcement learning (e.g., amygdala, ventral striatum, OFC/vlPFC) and attentional/control processes (e.g., dACC, pMCC, lateral PFC), offerring partial support for both accounts. However, the emotion- and cognitive-based theories cannot fully account for the robust effects found in other brain regions including the brainstem/midbrain, posterior hippocampus, superior temporal sulcus and visual cortices.

One of the earliest psychobiological theory of criminal and antisocial behaviors postulated that low cortical arousal would result in impaired behavioral conditioning, therefore increasing the proneness to stimulation seeking behaviors ^10^. Despite that it has long been proposed that low arousal would be associated with *fearlessness* in antisocial population ^9, 115, 116^, initial theories of personality rather linked low arousal to a mode of functioning primarily found in extraverted individuals ^117^, notably those with exaggerated reward seeking ^6^. According to this model, these individuals with low arousal would exhibit antisocial behaviors to produce shifts in motivational states. Coherently with this model, low arousal has been robustly linked to antisocial population including adolescents with elevated CP (and CD) ^20, 118, 119^, antisocial behaviors ^120^, psychopathy ^18^ and criminal offending, especially violent behaviors ^121, 122^. Not only this arousal theory can provide additional neurobiological insights to CD symptoms but may also link conceptually both emotion- and cognitive accounts. Indeed, autonomic arousal is fundamental for many cognitive processes including attention, memory, language, and executive functions ^123, 124^ as well as incentive learning ^125^. According to the Yerkes-Dobson Law, too much or too less arousal would alter cognitive performance. On a neurobiological level, Eysenck ^117^ already described the importance of the reticular formation ^126^, now known as the Ascending Arousal System, which is characterized by multiple neurotransmitter-specific projections (cholinergic, noradrenergic, dopaminergic, serotoninergic) from brainstem nuclei to the thalamus (dorsal pathway) and hypothalamus and basal forebrain (ventral pathway) ^62, 127^. This ascending arousal system further contribute to the coordination of cortical activity in many brain regions overlapping with those found in the CD-Network including the ACC, MCC, and fronto-insular cortex^128^.

While the neurobiological correlates of arousal appears to overlap with those found in CD, it may not be sufficient to explain widespread deficits in emotion perception such as emotion recognition and emotional resonance. Indeed, findings also suggest that CD youths have particularly lower fixation to the eyes when processing negative facial expressions ^97, 129–131^ and deficits in emotional resonance ^91, 132^. While many researchers adopt an amygdala-centric view of the neural dysfunctions in youths with CD, evidence indicates “*the amygdala is not essential for rapid, non-conscious detection of affective information.*” (p.6, ^133^). For example, visual information about emotional stimuli appears to be processed by pulvinar/lateral geniculate, visual cortices (V1, V2) and inferotemporal cortex as fast as 60-85ms, followed by 100-200 ms for the amygdala ^133^. This visual processing is often described via a two-pathway model ^134^, including the dorsal (“Where”) and ventral (“What”) streams. However, Pitcher & Ungerleider ^135^ recently suggested the presence of a third visual pathway specific to social perception (e.g., calculating meanings and intentions of others), which relies on projections from early visual cortex to the anterior portion of the superior temporal sulcus via the posterior superior temporal sulcus ^135^. These are largely overlapping with brain regions found in the CD-Network, spanning from the angular gyrus to the anterior superior temporal sulcus. Of importance, our perceptions are known to be shaped by the contextual information we gather when encountering particular stimuli (e.g., seeing other’s in distress). Evidence suggests that the retrieval of contextual representations is processed by the posterior hippocampus via the visual pathways and parahippocampal gyrus ^136, 137^, which were among the brain regions found to be the mostly replicated using the CD-network approach. In sum, it is thus possible that the neurobiological deficits regarding the perception of social cues may appear earlier in the temporal sequence than was previously assumed, as they involve both the perception and retrieval of contextual representations. Indeed, using a large sample of adolescents (n=1,416), we previously showed that severity of CP was significantly associated with functional connectivity of many brain regions found in the current study, providing additionnal evidence for a role of visual cortex, posterior hippocampus, angular gyrus, posterior temporal gyrus in CD ^138^. More research is needed to examine how the retrieval and integration of prior contextual knowledge may shape perception of social cues in youths with CD. Of importance, it is reported that only a limited number of youths with CD show poor emotion recognition skills (about 23%, ^139^), disentangling the heterogeneity of youths with CD should be a central goal in future neuroimaging studies.

### Limitations

A few limitations need to be acknowledged. First, fMRI studies on CD differ on a wide range of methodological and clinical features. Despite our attempt to examine how these features may impact our results (e.g., resting-state, CU traits, ADHD comorbidity, medication), the relatively small sample size limits our ability to examine other features. Similarly, since CD it often described by Aggression and Rule-Breaking subdimensions, it remains unknown whether some of our findings may be driven by one of these dimensions. Another limitation is the use of normative sample to characterize alterations in neural networks of CD. While normative approaches are powerful method to identify impairments, it is possible that youths with CD may show reorganization of the brain connectome ^140^ which may differ from normative expectations. Using mega-analytic approaches, such as in the ENIGMA-ASB Working Group, may help clarify to what extent CD youths show a brain reorganization compared to controls, respectively to our CD-Network.

## Conclusion

Researchers often describe the neural correlates of CD using a limited set of brain regions, noticeably the amygdala and prefrontal regions. However, neuroimaging literature reveal a more complex story than originally assumed by showing very weak spatial convergence across fMRI studies using a traditional meta-analytical approach. Here, we demonstrated that the heterogeneous peaks across these studies are connected to an overarching circuit. This distributed network, labelled as the CD-Network, mainly overlaps with neurobiological correlates of arousal-motivational and socio-affective processes which echo with prior work highlighting the importance of autonomic under-arousal and emotional perception in the emergence of antisocial behaviors. This CD-Network may serve as a template to study inter-individual differences in the neural processes that increase the risk for CD.

## Data Availability

All data produced in the present study are available upon reasonable request to the authors

## Acknowledgements

SP is holder of the Eli Lilly Canada Chair on schizophrenia research. JRD is holder of a postdoctoral fellowship from the Canadian Institutes of Health Research (MFE-181885).

## Authorship

Both authors have made substantial contributions to this work. JRD & SP conceptualized the study and interpreted the results. JRD did the statistical analyses and wrote the first draft of the manuscript. SP provided significant revision, and both authors approved the final version of this manuscript.

