## Supplementary Material for "Mapping the Brain Network of Conduct Disorder: Heterogeneous fMRI findings converge on a Common Brain Circuit"

### Supplementary Method

Activation Likelihood Estimation

Experiments' coordinates were used for spatial convergence using the Activation Likelihood Estimate method (GingerALE version 3.0.2,[1, 2]; http://www.brainmap.org/ale/). For each experiment, a 3D gaussian probability distribution was modelled around each coordinate foci, weighted by the number of subjects in the experiment. This method is performed to account for spatial uncertainty due to template and between-subject variance [1, 2]. It also ensures that multiple coordinates from a single experiment does not jointly influence the modeled activation value of a single voxel. The probabilities of all activation foci in an experiment were then combined. Voxel-wise ALE scores arise from the union across all these experiment maps. Consequently, a cluster-level corrected threshold was applied on the voxel-wise image. The size of the supra-threshold clusters was compared against a null distribution of cluster sizes derived from simulation of datasets. We used the following statistical threshold: $p<0.001$ at voxel-level and FWE-$p<0.05$ at a cluster-level with 5000 permutations.

Data Preprocessing

Normative functional connectivity map was conducted on data of the Autism Brain Imaging Data Exchange datasets, ABIDE-I [3, 4] and ABIDE-II [5]. Only data from >6 to <18 years old were preprocessed. A total of 687 subjects were preprocessed. Functional images were realigned, corrected for motion artifacts with the Artifact Detection Tool [6] (ART, setting a threshold of 0.9 mm subject ART's composite motion and a global signal threshold of Z = 5) with the implemented in CONN Toolbox [7], and co-registered to the corresponding anatomical image. The anatomical images were segmented (into grey matter, white matter, and cerebrospinal fluid) and normalized to the Montreal Neurological Institute (MNI) stereotaxic space. Functional images were then normalized based on structural data, spatially smoothed with a 6 mm full-width-at-half-maximum (FWHM) 3D isotropic Gaussian kernel and resampled to 2 $mm^3$ voxels. Bandpass filter was applied (0.01 Hz < f < 0.10 Hz). For the preprocessing, the anatomical component-based noise correction method (aCompCor strategy, [8]), was employed to remove confounding effects from the BOLD time series, such as the physiological noise originating from the white matter and cerebrospinal fluid. This method was found to increase the validity and sensitivity of analyses [9]. Preprocessed images were manually checked. We excluded 89 subjects which given their movement (e.g., average framewise displacement >.20 and/or percentage of scrubbed scans lower than 75%), leaving a remaining sample of 598 subjects.

### Supplementary Results

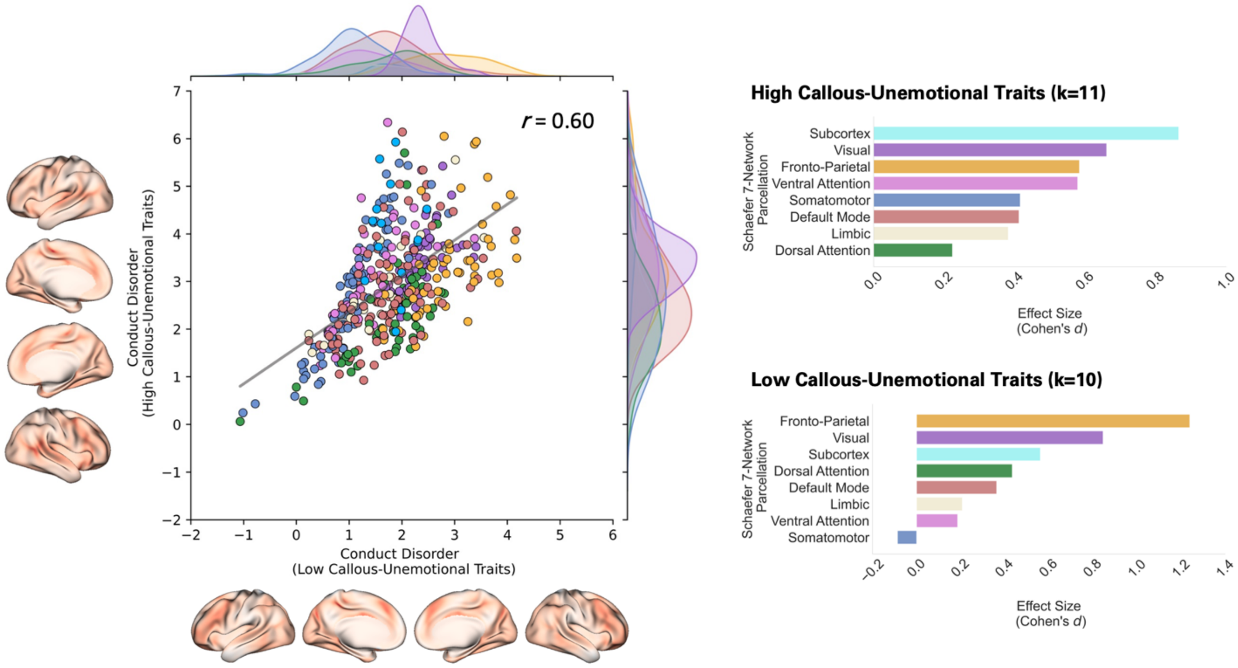

**Supplementary Figure 1.** Subanalyses on the effect of Callous-Unemotional Traits using a Median-Split approach. Of the 38 identified studies, only 21 reported their levels of CU traits, which were further grouped into high (k=11, POMP-CU=.68) and low (k=10, POMP-CU=.43) using percentage of maximum possible score.

**Supplementary Table 1.** Gene-Category Enrichment Analyses of the CD-Network (DisGeNet)

| Phenotypes | Number of Genes | Spearman rho (r-to-z) | p-values (uncorrected) | p-values (corrected) | p-values (permutation +uncorrected) | p-values (permutation+ corrected) | Standardized Weighted Correlation |
|---|---|---|---|---|---|---|---|
| Psychosocial stressor | 11 | 0.107 | 1.3E-07 | 2.7E-05 | 0.0E+00 | 0.0E+00 | 3.22 |
| Sexual inhibition | 2 | 0.226 | 1.1E-07 | 2.7E-05 | 0.0E+00 | 0.0E+00 | 2.74 |
| Impaired ability to form peer relationships | 2 | 0.216 | 3.2E-07 | 5.0E-05 | 0.0E+00 | 0.0E+00 | 2.44 |
| Childhood disintegrative disorder | 2 | 0.207 | 2.3E-06 | 2.4E-04 | 0.0E+00 | 0.0E+00 | 2.17 |
| Psychological pseudocyesis | 8 | 0.103 | 3.7E-06 | 3.3E-04 | 0.0E+00 | 0.0E+00 | 2.16 |
| Pathological personality NOS | 3 | 0.157 | 4.7E-06 | 3.6E-04 | 0.0E+00 | 0.0E+00 | 1.92 |
| Hypersexuality state | 6 | 0.104 | 2.2E-05 | 9.6E-04 | 0.0E+00 | 0.0E+00 | 1.50 |
| Cocaine withdrawal | 10 | 0.078 | 1.6E-05 | 8.1E-04 | 0.0E+00 | 0.0E+00 | 1.09 |
| psychiatric hospitalization | 3 | 0.133 | 7.9E-05 | 2.3E-03 | 2.0E-04 | 4.5E-03 | 1.04 |
| Atypical autism | 3 | 0.131 | 9.2E-06 | 6.3E-04 | 0.0E+00 | 0.0E+00 | 0.97 |
| Major depression, single episode | 32 | 0.051 | 4.5E-08 | 2.7E-05 | 0.0E+00 | 0.0E+00 | 0.86 |
| Cluster C personality disorder | 2 | 0.154 | 1.3E-04 | 3.4E-03 | 4.0E-04 | 7.4E-03 | 0.63 |
| Neurotic personality | 2 | 0.154 | 1.3E-04 | 3.4E-03 | 4.0E-04 | 7.4E-03 | 0.63 |
| Severe expressive language delay | 11 | 0.068 | 4.4E-04 | 8.1E-03 | 6.0E-04 | 9.7E-03 | 0.59 |
| Restrictive behavior | 12 | 0.065 | 5.3E-05 | 1.9E-03 | 0.0E+00 | 0.0E+00 | 0.57 |
| Paranoid Schizophrenia | 34 | 0.047 | 5.2E-05 | 1.9E-03 | 2.0E-04 | 4.5E-03 | 0.54 |
| Auditory and visual hallucinations | 2 | 0.151 | 1.4E-05 | 7.8E-04 | 0.0E+00 | 0.0E+00 | 0.54 |
| Seasonal Affective Disorder | 42 | 0.043 | 9.0E-07 | 1.1E-04 | 0.0E+00 | 0.0E+00 | 0.47 |
| Depression and Suicide | 9 | 0.071 | 3.9E-03 | 3.1E-02 | 4.6E-03 | 3.6E-02 | 0.45 |
| Memory, Short-Term | 8 | 0.074 | 5.4E-04 | 9.2E-03 | 4.0E-04 | 7.4E-03 | 0.44 |
| Eating disorder symptom | 11 | 0.064 | 2.3E-04 | 5.4E-03 | 2.0E-04 | 4.5E-03 | 0.35 |
| Adolescent antisocial behaviour | 2 | 0.144 | 3.3E-04 | 7.0E-03 | 4.0E-04 | 7.4E-03 | 0.34 |
| Mania acute | 2 | 0.144 | 1.1E-05 | 6.5E-04 | 0.0E+00 | 0.0E+00 | 0.32 |
| Stereotypical body rocking | 3 | 0.114 | 4.0E-04 | 8.0E-03 | 2.0E-04 | 4.5E-03 | 0.32 |
| Processing speed | 4 | 0.098 | 7.6E-05 | 2.3E-03 | 2.0E-04 | 4.5E-03 | 0.31 |
| Premenstrual Dysphoric Disorder | 8 | 0.070 | 4.2E-04 | 8.0E-03 | 4.0E-04 | 7.4E-03 | 0.21 |
| Mild dementia | 10 | 0.063 | 3.1E-04 | 7.0E-03 | 1.0E-03 | 1.5E-02 | 0.16 |
| Cyclothymic Disorder | 2 | 0.133 | 2.0E-05 | 9.6E-04 | 0.0E+00 | 0.0E+00 | 0.01 |
| Impaired use of nonverbal behaviors | 6 | 0.075 | 2.9E-05 | 1.2E-03 | 0.0E+00 | 0.0E+00 | -0.02 |
| Violence | 46 | 0.038 | 1.0E-03 | 1.7E-02 | 1.4E-03 | 1.9E-02 | -0.02 |
| Heroin Smoking | 6 | 0.075 | 2.7E-03 | 2.5E-02 | 3.8E-03 | 3.2E-02 | -0.02 |
| Emotional Disturbances | 5 | 0.079 | 2.4E-03 | 2.3E-02 | 2.8E-03 | 2.6E-02 | -0.10 |
| Specific reading disorder | 3 | 0.102 | 1.1E-03 | 1.8E-02 | 8.0E-04 | 1.3E-02 | -0.11 |
| Alexia | 9 | 0.060 | 1.4E-03 | 2.1E-02 | 1.4E-03 | 1.9E-02 | -0.19 |
| Catatonia | 12 | 0.053 | 4.2E-03 | 3.2E-02 | 4.4E-03 | 3.5E-02 | -0.24 |
| Severe depression | 25 | 0.042 | 1.1E-04 | 3.0E-03 | 0.0E+00 | 0.0E+00 | -0.25 |
| Euthymia | 5 | 0.076 | 7.2E-05 | 2.3E-03 | 0.0E+00 | 0.0E+00 | -0.26 |
| Alcohol problem | 18 | 0.046 | 1.8E-03 | 2.3E-02 | 1.6E-03 | 2.1E-02 | -0.27 |
| Shyness | 9 | 0.059 | 3.6E-03 | 3.0E-02 | 4.4E-03 | 3.5E-02 | -0.29 |
| Oral aversion | 7 | 0.064 | 4.6E-04 | 8.3E-03 | 6.0E-04 | 9.7E-03 | -0.33 |
| Alexithymia | 28 | 0.039 | 1.7E-03 | 2.3E-02 | 1.4E-03 | 1.9E-02 | -0.39 |
| Mental Retardation (X-Linked, 89) | 3 | 0.095 | 2.2E-03 | 2.3E-02 | 1.2E-03 | 1.8E-02 | -0.40 |
| Minimal Brain Dysfunction | 18 | 0.044 | 3.4E-03 | 3.0E-02 | 3.6E-03 | 3.1E-02 | -0.41 |

| | | | | | | | |
|---|---|---|---|---|---|---|---|
| Spoken Word Recognition Deficit | 8 | 0.059 | 2.1E-03 | 2.3E-02 | 2.6E-03 | 2.5E-02 | -0.44 |
| Impulsive character (finding) | 32 | 0.036 | 3.7E-03 | 3.0E-02 | 4.4E-03 | 3.5E-02 | -0.54 |
| Cluster B personality disorder | 3 | 0.091 | 3.4E-03 | 3.0E-02 | 4.2E-03 | 3.5E-02 | -0.54 |
| Executive dysfunction | 27 | 0.038 | 5.1E-04 | 8.9E-03 | 2.0E-04 | 4.5E-03 | -0.56 |
| Fear of heights | 2 | 0.113 | 1.4E-03 | 2.1E-02 | 1.4E-03 | 1.9E-02 | -0.60 |
| Abnormal fear/anxiety-related behavior | 5 | 0.069 | 4.2E-03 | 3.2E-02 | 5.2E-03 | 3.9E-02 | -0.60 |
| Aloof | 61 | 0.030 | 1.3E-03 | 1.9E-02 | 6.0E-04 | 9.7E-03 | -0.61 |
| Depersonalization | 3 | 0.089 | 6.5E-05 | 2.2E-03 | 0.0E+00 | 0.0E+00 | -0.62 |
| heroin abuse | 14 | 0.045 | 3.1E-03 | 2.9E-02 | 4.2E-03 | 3.5E-02 | -0.64 |
| Social Anxiety | 44 | 0.032 | 1.6E-04 | 3.9E-03 | 2.0E-04 | 4.5E-03 | -0.68 |
| Compulsive sexual behaviour | 2 | 0.110 | 1.8E-03 | 2.3E-02 | 1.8E-03 | 2.4E-02 | -0.68 |
| Other and unspecified reactive psychosis | 6 | 0.062 | 3.2E-03 | 2.9E-02 | 2.0E-03 | 2.5E-02 | -0.69 |
| Impulsive aggression | 10 | 0.049 | 3.9E-03 | 3.1E-02 | 5.4E-03 | 4.0E-02 | -0.73 |
| Ritual compulsion | 7 | 0.057 | 5.2E-03 | 3.9E-02 | 6.0E-03 | 4.4E-02 | -0.75 |
| Bulimia Nervosa | 47 | 0.030 | 5.3E-03 | 3.9E-02 | 6.0E-03 | 4.4E-02 | -0.76 |
| Polydrug abuse | 2 | 0.105 | 5.8E-03 | 4.2E-02 | 5.2E-03 | 3.9E-02 | -0.82 |
| Physical violence | 3 | 0.082 | 4.3E-03 | 3.2E-02 | 3.6E-03 | 3.1E-02 | -0.85 |
| Antisocial Personality Disorder | 44 | 0.030 | 1.9E-03 | 2.3E-02 | 3.0E-03 | 2.7E-02 | -0.89 |
| Negative affectivity | 8 | 0.051 | 4.1E-04 | 8.0E-03 | 6.0E-04 | 9.7E-03 | -0.90 |
| Emotional Stress | 17 | 0.039 | 3.3E-04 | 7.0E-03 | 2.0E-04 | 4.5E-03 | -0.91 |
| Derealization | 2 | 0.102 | 1.7E-03 | 2.3E-02 | 6.0E-04 | 9.7E-03 | -0.92 |
| Emotional neglect | 5 | 0.062 | 3.6E-03 | 3.0E-02 | 3.4E-03 | 3.0E-02 | -0.96 |
| Aphasia, Progressive | 8 | 0.049 | 6.2E-04 | 1.0E-02 | 4.0E-04 | 7.4E-03 | -1.00 |
| Other eating disorders | 5 | 0.057 | 3.5E-03 | 3.0E-02 | 3.2E-03 | 2.9E-02 | -1.16 |
| Diagnosis, Psychiatric | 32 | 0.028 | 2.5E-03 | 2.4E-02 | 2.8E-03 | 2.6E-02 | -1.28 |
| Aphasia, Acquired | 2 | 0.089 | 2.4E-03 | 2.3E-02 | 2.6E-03 | 2.5E-02 | -1.30 |
| Chronic schizophrenia | 38 | 0.024 | 3.3E-03 | 2.9E-02 | 2.6E-03 | 2.5E-02 | -1.58 |
| Moderate expressive language delay | 2 | 0.076 | 5.4E-03 | 4.0E-02 | 5.2E-03 | 3.9E-02 | -1.68 |

Note. Only Phenotypes reaching p<0.05 after correction with 5,000 permutations are displayed

**Supplementary Table 2.** Identified Genes and gene-wise correlations with CD-Network

| Genes | Rates of Occurrence (%) | Spearman Rho | p-values |
|---|---|---|---|
| BDNF | 39.44% | 0.159 | 2.56E-03 |
| MAOA | 28.17% | 0.192 | 2.47E-04 |
| DRD2 | 25.35% | 0.095 | 7.14E-02 |
| COMT | 19.72% | 0.021 | 6.93E-01 |
| HTR1A | 19.72% | 0.148 | 5.03E-03 |
| HTR2A | 18.31% | 0.025 | 6.37E-01 |
| DRD4 | 15.49% | -0.013 | 8.02E-01 |
| OXTR | 15.49% | 0.120 | 2.29E-02 |
| HTR2C | 12.68% | 0.308 | 2.62E-09 |
| TNF | 12.68% | -0.071 | 1.80E-01 |
| POMC | 11.27% | 0.106 | 4.54E-02 |
| SLC6A3 | 11.27% | 0.044 | 4.09E-01 |
| GABRA2 | 9.86% | 0.116 | 2.85E-02 |
| BAG3 | 8.45% | 0.037 | 4.89E-01 |
| C9orf72 | 8.45% | 0.015 | 7.83E-01 |
| GRN | 8.45% | 0.097 | 6.50E-02 |
| NOS1 | 8.45% | 0.146 | 5.69E-03 |
| ACE | 7.04% | -0.092 | 8.29E-02 |
| APOE | 7.04% | 0.046 | 3.85E-01 |
| ATF7IP | 7.04% | 0.083 | 1.14E-01 |
| FKBP5 | 7.04% | 0.107 | 4.20E-02 |
| PSEN1 | 7.04% | 0.061 | 2.45E-01 |
| TAL1 | 7.04% | 0.042 | 4.25E-01 |
| ANKK1 | 5.63% | -0.017 | 7.48E-01 |
| ARSD | 5.63% | -0.046 | 3.83E-01 |
| AVP | 5.63% | 0.015 | 7.71E-01 |
| CNR1 | 5.63% | 0.079 | 1.37E-01 |
| CRH | 5.63% | -0.101 | 5.55E-02 |
| EBPL | 5.63% | -0.098 | 6.26E-02 |
| ELK3 | 5.63% | 0.071 | 1.77E-01 |
| ESR1 | 5.63% | 0.036 | 5.02E-01 |
| FAAH | 5.63% | 0.198 | 1.57E-04 |
| GAD1 | 5.63% | 0.023 | 6.64E-01 |
| IL1B | 5.63% | -0.013 | 8.06E-01 |
| MAOB | 5.63% | 0.176 | 8.23E-04 |
| MECP2 | 5.63% | 0.001 | 9.92E-01 |
| NLGN4X | 5.63% | 0.313 | 1.27E-09 |
| NR3C1 | 5.63% | -0.174 | 9.59E-04 |
| NTRK2 | 5.63% | -0.023 | 6.67E-01 |
| OPRM1 | 5.63% | -0.094 | 7.68E-02 |
| SNCA | 5.63% | 0.001 | 9.84E-01 |
| WASF1 | 5.63% | 0.121 | 2.19E-02 |

Note. Only Genes >5% Rates of Occurrence are displayed
